# MyoScore: a Genetically Anchored Transcriptomic Scoring System for Quantifying Human Skeletal Muscle Health

**DOI:** 10.64898/2026.05.11.26352204

**Authors:** Huahua Zhong, Shang Ma, Victoria Lillback, Mingshi Gao, Wenhan Zhang, Mridul Johari, Ali Oghabian, Per Harald Jonson, Jianying Xi, Wenhua Zhu, Shanfeng Zhu, Peter Hackman, Bjarne Udd, Chongbo Zhao, Marco Savarese, Sushan Luo

## Abstract

Muscle health varies continuously from optimal function to severe pathology, yet no unified genetic framework quantifies this spectrum objectively. Here we develop MyoScore, a transcriptomic scoring system derived from transcriptome-wide association studies of 27 muscle-related phenotypes in over one million participants. TWAS selects genes whose genetically regulated expression in skeletal muscle associates with muscle-related traits, providing the genetic anchoring of the scoring system, while MyoScore itself is computed from measured bulk RNA-seq expression in new samples. From 1,116 transcriptome-wide association study (TWAS)-significant genes, 417 are expressed in skeletal muscle and form the basis of the scoring system. These genes are organized into five dimensions of muscle biology (Strength, Mass, LeanMuscle, Youth and Resilience), each scored from 0 to 100. Across 1,722 human skeletal muscle transcriptomes from four independent cohorts, MyoScore defines a continuous four-stage muscle health spectrum, discriminates healthy from diseased muscle (area under the curve 0.751–0.873), and correlates with histopathological severity, quantitative MRI and clinical outcomes. Functional validation through iPSC-to-myotube differentiation supports predicted expression changes for novel MyoScore genes. UK Biobank analysis of blood biomarker proxies in 467,123 participants demonstrates concordant associations with muscle phenotypes, and two-sample Mendelian randomization using skeletal muscle cis-eQTL supports causal directionality for 78% of gene-outcome pairs tested. Single-cell validation across 475,584 cells from two independent muscle ageing atlases shows that pseudobulk MyoScore declines with age, with type II myofibre nuclei most affected. Together, MyoScore establishes the first genetically anchored, dimension-resolved quantification of human muscle health, enabling objective assessment, patient stratification and biomarker discovery across the full spectrum from optimal function to severe disease.

## Introduction

Skeletal muscle accounts for approximately 40% of body mass and is the primary site of insulin-stimulated glucose disposal^1^, making it a central organ in whole-body metabolic homeostasis. Skeletal muscle diseases encompass a broad clinical spectrum, from age-related sarcopenia affecting 10–27% of elderly populations^2^ to severe hereditary myopathies with prevalences of 3.6–7.1 per 100,000^3,4^. Aging-related muscle decline, now recognized as a spectrum extending beyond traditional sarcopenia thresholds, contributes substantially to frailty, falls and loss of independence in the elderly, and is increasingly implicated as a co-morbidity in neurodegenerative disease and cancer cachexia^5^. Despite this heterogeneity, clinical assessment remains limited to semi-quantitative functional scales, invasive biopsies with qualitative histopathological evaluation, muscle MRI interpretation that retains substantial inter-reader variability, and non-specific biomarkers such as serum creatine kinase^6^. Transcriptomic studies have identified molecular signatures of individual myopathies^7,8^, including our previous characterization of expression patterns across the muscle health spectrum^9^, but no unified framework exists to quantify muscle health objectively across the complete continuum from healthy to severely diseased states.

Large-scale genome-wide association studies (GWAS), particularly from the UK Biobank^10^, have revealed the polygenic architecture of muscle-related traits, including grip strength^11^, lean body mass^12^, and MRI-derived fat infiltration measures^13^. However, variant-level associations provide limited biological insight, as they do not capture the tissue-specific gene expression effects that ultimately determine phenotypes^14^. Transcriptome-wide association studies (TWAS) offer a powerful approach to bridge this gap by integrating GWAS summary statistics with expression quantitative trait loci (eQTL) from relevant tissues^15,16^. The FUSION framework, leveraging GTEx v8 skeletal muscle eQTL weights from 803 samples^17^, enables systematic identification of genes whose genetically regulated expression influences muscle-related traits.

Muscle health is inherently multidimensional, encompassing functional capacity, tissue composition, biological ageing and disease resistance^18^. While multi-trait genetic approaches have successfully identified shared architectures in psychiatric and cardiometabolic disorders^19,20^, no systematic integration of diverse muscle GWAS phenotypes into a unified framework has been attempted. Here we present MyoScore, a genetically anchored transcriptomic scoring system that integrates TWAS results from 27 muscle-related GWAS phenotypes to quantify muscle health across five biologically interpretable dimensions. The “genetic anchoring” refers to the fact that both the identity of scoring genes and the direction in which each gene’s expression is weighted are derived from TWAS genetic evidence (GWAS summary statistics integrated with skeletal muscle cis-eQTL), rather than from phenotype-driven differential expression; the resulting score is then computed from conventional bulk RNA-seq expression in each sample. We validate MyoScore across 1,722 human skeletal muscle transcriptomes from four independent cohorts, demonstrate concordance with histopathological, imaging and clinical measures, functionally validate novel genes through iPSC differentiation, support causal directionality through Mendelian randomization with tissue-matched instruments, identify blood biomarker proxies in up to 467,123 UK Biobank participants, and confirm single-cell validity across two independent muscle ageing atlases.

## Results

### TWAS integration yields a five-dimensional muscle health framework

To construct a comprehensive genetic framework for muscle health, we performed TWAS across 27 muscle-related phenotypes from large-scale GWAS studies encompassing over 1,000,000 participants (Fig. 1). Phenotypes were grouped into five biologically interpretable domains reflecting distinct aspects of muscle function and pathology: functional capacity, anthropometric tissue composition, MRI-derived tissue quality, biological ageing and disease phenotype. Phenotypes spanned functional measures (grip strength, walking pace), body composition (lean mass, fat-free mass), MRI-derived tissue characteristics (thigh fat infiltration), biological ageing (telomere length) and disease outcomes (muscular dystrophy, myopathies, serum creatine kinase levels). Using the FUSION framework^16^ with GTEx v8 skeletal muscle eQTL weights^17^, we tested associations for 2,975 genes with significant cis-heritable expression.

**Fig. 1.**
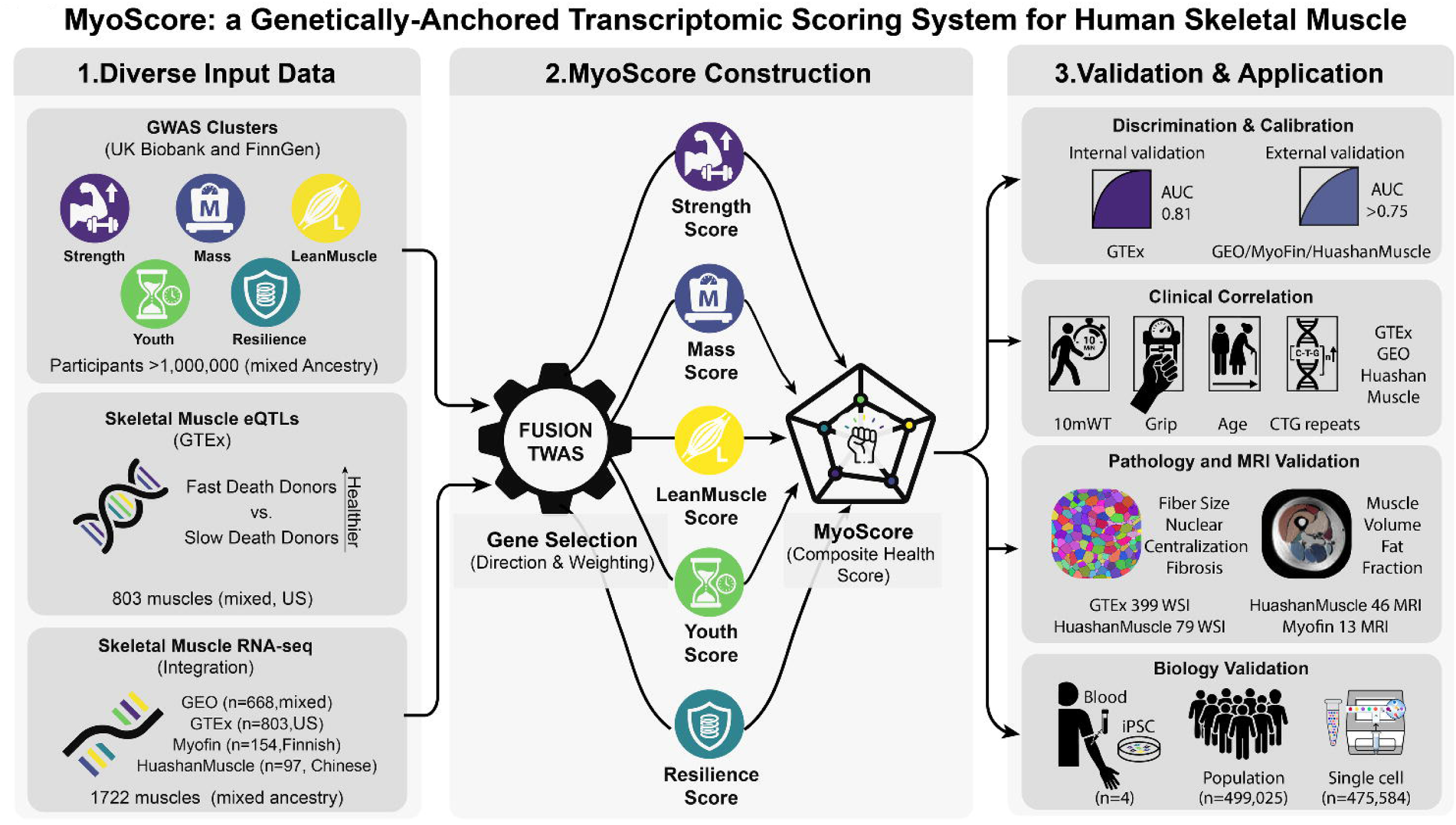
Development of the five-dimensional MyoScore framework. Overview of the TWAS pipeline: 27 muscle-related GWAS phenotypes from over 1 million participants are integrated with GTEx v8 skeletal muscle eQTL weights (n = 803) through FUSION TWAS, identifying 1,116 significant genes. Organization of genes into five dimensions (Strength, Mass, LeanMuscle, Youth, Resilience) with gene counts and data-driven weights. MyoScore calculation workflow: gene-wise Z-score normalization, weighted averaging within dimensions, min–max scaling to 0–100 and composite scoring.

After Bonferroni correction (P < 1.68 × 10^−5^), we identified 1,116 genes with significant TWAS associations. Of these, 591 gene–dimension entries corresponding to 417 unique skeletal-muscle-expressed genes were detectable in bulk RNA-seq expression matrices and formed the basis of MyoScore calculation. Because individual genes can be significant in GWAS phenotypes assigned to different dimensions (reflecting biological pleiotropy), each gene was counted once per dimension in which it reached significance; dimension sub-scores are computed independently over their respective gene sets and subsequently combined, so pleiotropic genes contribute proportionally to each dimension on which they are TWAS-significant rather than being double-counted inadvertently. We organized these skeletal-muscle-expressed genes into five dimensions based on the physiological relevance of their source phenotypes: Strength (67 total, 31 detectable genes from grip strength and walking pace), Mass (382 total, 219 detectable from 15 lean body mass phenotypes), LeanMuscle (272 total, 147 detectable from MRI fat infiltration, direction reversed so that higher scores indicate less fat), Youth (81 total, 37 detectable from telomere length GWAS) and Resilience (314 total, 157 detectable from myopathy diagnoses and creatine kinase, direction reversed so that higher scores indicate greater disease resistance).

The Youth dimension captures biological ageing through telomere length GWAS^21^. The 81 Youth genes include established telomere biology factors: OBFC1 (STN1), a CST complex component essential for telomere maintenance^22,23^; TERF2, a shelterin complex member^24^; and NAF1, required for telomerase assembly^25^. This enrichment supports the biological relevance of this dimension. Gene weights were determined by −log^10^(P value) from TWAS, and effect directions were assigned from TWAS Z-scores with reversals applied to negative phenotypes to ensure that higher expression of positively weighted genes consistently indicates better muscle health. Dimension sub-scores were combined into a composite MyoScore using data-driven weights derived on the GTEx reference cohort (Supplementary Note 1): MyoScore = 0.252 × Strength + 0.177 × Mass + 0.243 × LeanMuscle + 0.242 × Youth + 0.087 × Resilience.

### Pathway analysis reveals metabolic functions of MyoScore dimensions

To assess whether the genetically identified dimensions capture coherent biological functions, we performed gene ontology (GO) and KEGG pathway enrichment analysis for each dimension (Fig. 2). The Strength dimension showed the most pronounced metabolic enrichment, with significant overrepresentation of acetyl-CoA metabolic processes including acid-thiol ligase and CoA-ligase activity (adjusted P = 0.023; *ACSS2*, *ACSS3*), ethanol metabolic process (adjusted P = 0.004; *SULT1A1*, *ACSS2*, *SULT1A2*) and propanoate metabolism (adjusted P = 0.049; *ACSS2*, *ACSS3*) (Fig. 2A). These pathways converge on the conversion of short-chain fatty acids to acetyl-CoA, a central node in skeletal muscle energy metabolism^26^. Notably, *ACSS2* and *ACSS3*(Fig. 2B), the two acetyl-CoA synthetases enriched in Strength, were among the genes subsequently validated through UK Biobank blood biomarker analysis (described below), establishing a direct mechanistic link between the genetic signal and metabolic function. Strength genes were additionally enriched for microtubule binding (adjusted P = 0.023; *MAP1LC3A*, *KIF1B*, *MAPT*, *LRPPRC*), mitochondrial transport along microtubules (adjusted P = 0.047; *MAPT*, *LRPPRC*) and fibroblast growth factor binding (adjusted P = 0.030; *FGFR3*, *FGFRL1*), consistent with the cytoskeletal and growth-factor signalling requirements of functional muscle.

**Fig. 2.**
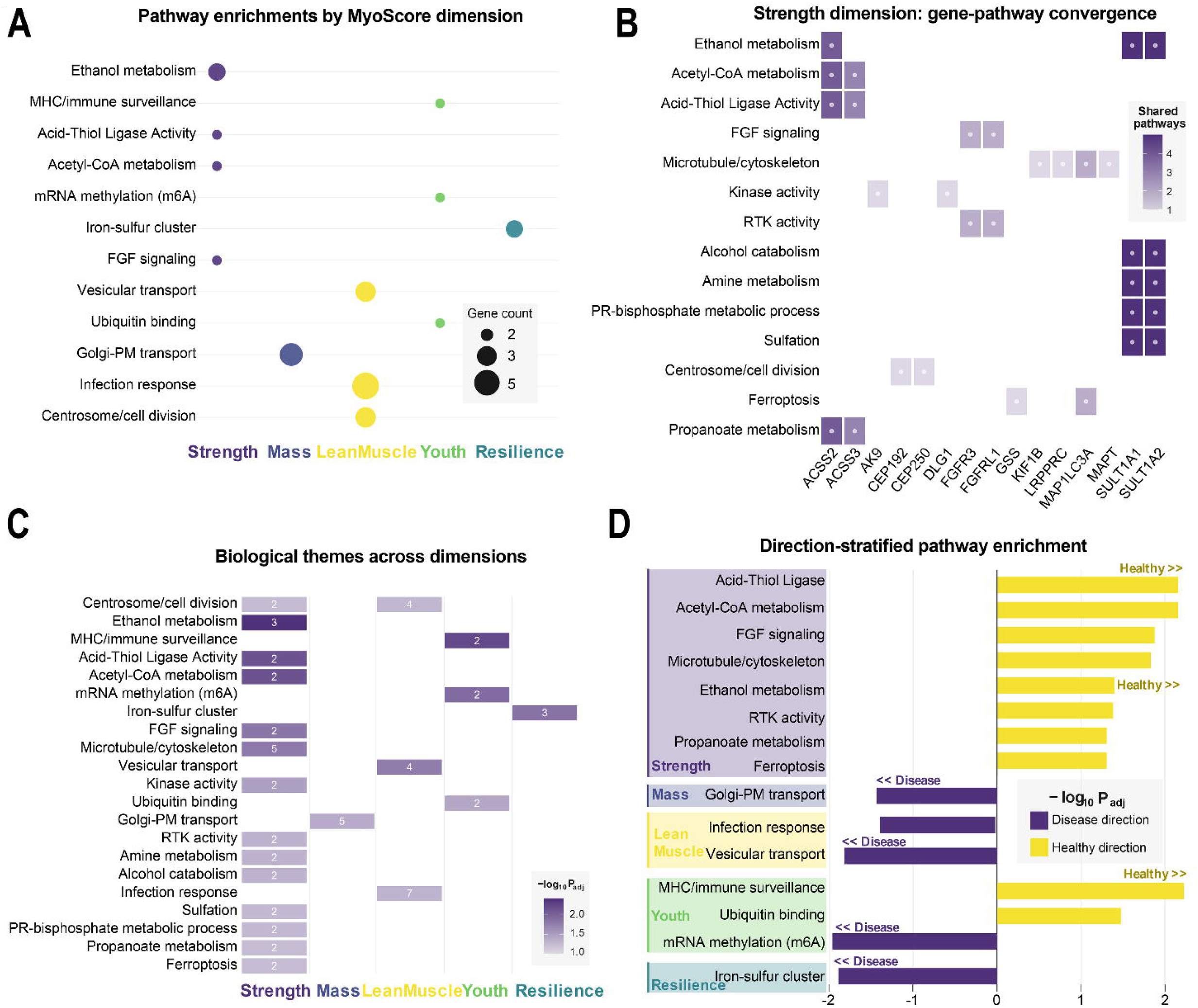
Pathway enrichment analysis reveals dimension-specific biological programmes underlying MyoScore. **a**, Bubble plot showing significantly enriched GO and KEGG pathways for each MyoScore dimension (adjusted P < 0.05). Bubble size reflects the number of enriched genes per pathway; bubble colour indicates dimension identity. **b**, Gene–pathway convergence matrix for the Strength dimension. Rows represent enriched biological themes; columns represent individual genes; colour intensity indicates the number of shared pathways (scale: 1–4). **c**, Heatmap of biological theme enrichment across all five dimensions. Bar length represents the number of significantly enriched gene sets per theme; colour intensity reflects enrichment significance (−log₁₀ adjusted P). **d**, Direction-stratified pathway enrichment for each dimension. Yellow bars indicate healthy-direction enrichment; purple bars indicate disease-direction enrichment. Significance threshold: Benjamini–Hochberg adjusted P < 0.05.

The LeanMuscle dimension was enriched for protein localization to the centrosome (adjusted P = 0.045; *NUDCD3*, *DCTN2*, *CEP250*, *CEP192*) (Fig. 2C), consistent with the role of centrosomal proteins in myoblast differentiation and post-mitotic cellular organization^27^. Direction-stratified analysis revealed that LeanMuscle genes whose upregulation indicates disease (unhealthy direction) were enriched in vesicular transport pathways including vasopressin-regulated water reabsorption (KEGG, adjusted P = 0.015; DYNC1I2, RAB5B, *DCTN2*, STX4), suggesting that disrupted intracellular trafficking contributes to fat infiltration in diseased muscle^28^.

The Youth dimension was enriched for mRNA methyltransferase activity (adjusted P = 0.037; METTL16, TRMT61B) and MHC class I protein binding (adjusted P = 0.037; PILRA, PILRB) (Fig. 2D). METTL16 catalyses N^6^-methyladenosine (m^6^A) modification of mRNA and regulates S-adenosylmethionine homeostasis^29^, linking biological ageing to epitranscriptomic regulation, a pathway implicated in age-related metabolic decline^30^. The MHC class I enrichment among Youth healthy-direction genes (adjusted P = 0.006) is consistent with previously reported declines in immune surveillance capacity with biological ageing in muscle tissue^31^.

The Resilience dimension showed enrichment for iron–sulfur cluster binding among disease-direction genes (adjusted P = 0.013; CISD1, CISD2, ISCU), implicating mitochondrial iron homeostasis and ferroptosis susceptibility in disease resistance^32^. CISD2 deficiency accelerates skeletal muscle ageing in mice^33^, and iron–sulfur cluster assembly is essential for respiratory chain function. The Mass dimension showed enrichment for Golgi-to-plasma membrane transport (adjusted P = 0.037), reflecting the secretory and membrane remodelling demands of muscle mass maintenance^34^.

Together, these enrichment patterns demonstrate that the five TWAS-derived dimensions capture distinct metabolic and cellular programmes: energy substrate metabolism (Strength), intracellular trafficking and tissue composition (LeanMuscle), epitranscriptomic ageing (Youth), mitochondrial iron homeostasis (Resilience) and membrane dynamics (Mass).

### MyoScore defines a continuous muscle health spectrum

Uniform manifold approximation and projection (UMAP) dimensionality reduction of all 1,722 skeletal muscle samples, overlaid with MyoScore values, revealed a continuous gradient from healthy to severely diseased muscle rather than discrete clusters (Fig. 3A). The UMAP embedding and MyoScore were computed from the same 417 skeletal-muscle-expressed TWAS genes. Diffusion component 1, capturing the dominant axis of transcriptomic variation defined on the same gene set, correlated significantly and in the expected direction with MyoScore (r = 0.417, P = 1.37 × 10^−73^), indicating that MyoScore is aligned with the primary biological axis of variation in muscle transcriptomes evaluated under this gene set.

**Fig. 3.**
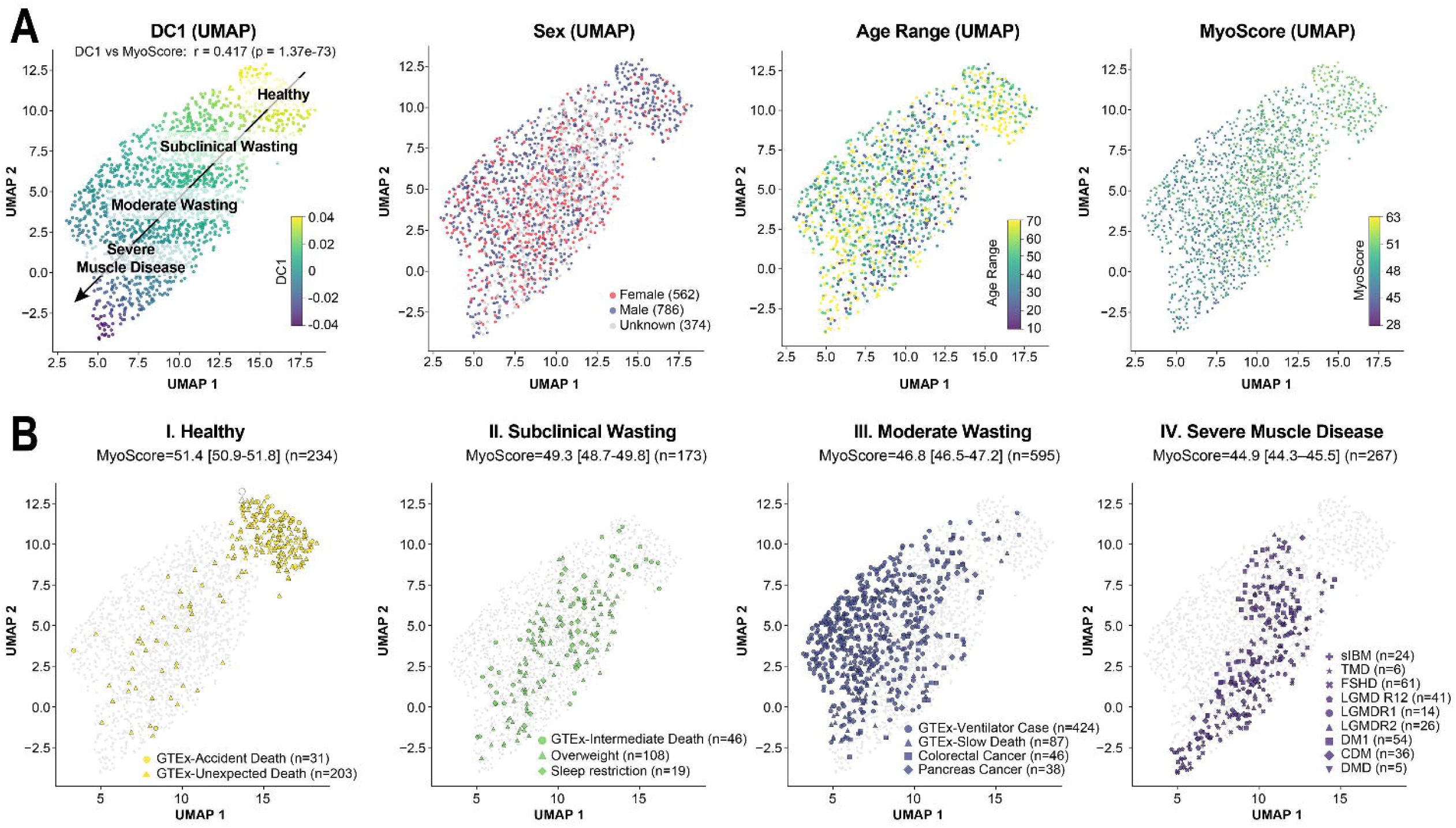
MyoScore defines a continuous muscle health spectrum. **a**, UMAP embedding of 1,722 skeletal muscle transcriptomes coloured by MyoScore (left) and diffusion component 1 (right). Pearson r = 0.417, P = 1.37 × 10^−73^. **b**, Four-stage muscle health spectrum defined by UMAP topology and MyoScore distributions. Error bars represent 95% CIs. White dots indicate individual samples not assigned to the highlighted stage.

On the basis of UMAP topology and MyoScore distributions, we defined a four-stage muscle health spectrum (Fig. 3B). Stage I (Healthy; MyoScore = 51.4, 95% confidence interval (CI) 50.9–51.8, n = 234) comprised GTEx donors with accidental or unexpected death. Stage II (Subclinical Wasting; MyoScore = 49.3, 95% CI 48.7–49.8, n = 173) included GTEx intermediate death donors, overweight individuals and sleep-restricted subjects. Stage III (Moderate Wasting; MyoScore = 46.8, 95% CI 46.5–47.2, n = 595) encompassed GTEx ventilator case and slow death donors alongside cancer-associated muscle wasting cohorts. Stage IV (Severe Muscle Disease; MyoScore = 44.9, 95% CI 44.3–45.5, n = 267) included hereditary myopathies: sporadic inclusion body myositis (sIBM), tibial muscular dystrophy (TMD), facioscapulohumeral dystrophy (FSHD), limb-girdle muscular dystrophy (LGMD) subtypes, myotonic dystrophy type 1 (DM1), congenital DM (CDM) and Duchenne muscular dystrophy (DMD). This classification demonstrates that muscle health decline extends continuously across chronic illness, cancer cachexia and hereditary myopathies.

### Technical robustness across platforms and conditions

We established MyoScore’s robustness through three complementary analyses. Paired samples (n = 3) sequenced on both DNBSEQ-T7 (MGI) and NovaSeq 6000 (Illumina) showed no statistically distinguishable difference in MyoScore (Wilcoxon test, P = 0.37) (Fig. 4A), supporting robustness across distinct sequencing chemistries. Paired samples (n = 12) prepared with polyA selection versus ribosomal depletion showed concordant scores (P = 0.29) (Fig. 4B). Within-individual muscle consistency, assessed using paired rectus femoris and vastus lateralis biopsies from 14 healthy individuals, demonstrated significant positive correlation (r = 0.60, P = 0.032) (Fig. 4C).

**Fig. 4.**
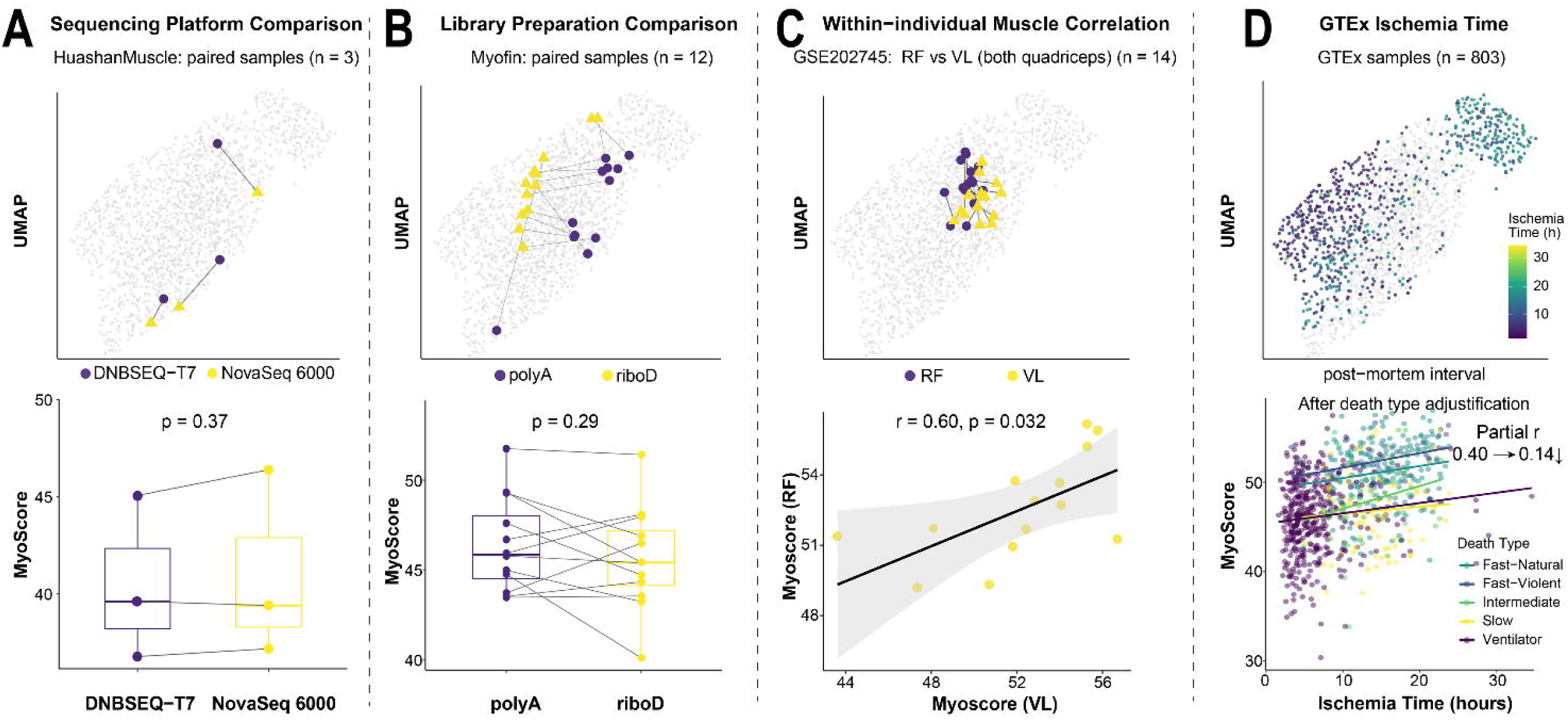
Technical robustness of MyoScore. **a**, Paired samples sequenced on DNBSEQ-T7 and NovaSeq 6000 (n = 3 pairs; Wilcoxon P = 0.37). **b**, Paired samples with polyA selection versus ribosomal depletion (n = 12 pairs; P = 0.29). **c**, Within-individual muscle consistency between rectus femoris and vastus lateralis (n = 14; r = 0.60, P = 0.032). **d**, GTEx ischemic time: raw r = 0.40, partial r (controlling for death type) = 0.14.

An apparent positive correlation between ischemic time and MyoScore (r = 0.40) in GTEx samples reflects confounding by death type rather than a genuine relationship between post-mortem interval and transcriptomic health: after adjusting for death type, the partial correlation reduced to 0.14 (Fig. 4D), supporting the interpretation that MyoScore reflects biological health status rather than post-mortem sample degradation.

### Disease discrimination and clinical validation

Receiver operating characteristic (ROC) analysis demonstrated consistent disease discrimination across all four cohorts (Fig. 5A): GTEx (area under the curve (AUC) = 0.825, 95% CI 0.793–0.855), GEO (AUC = 0.786, 95% CI 0.749–0.822), Helsinki Myofin (AUC = 0.751, 95% CI 0.588–0.886) and Shanghai HuashanMuscle (AUC = 0.873, 95% CI 0.786–0.948).

**Fig. 5.**
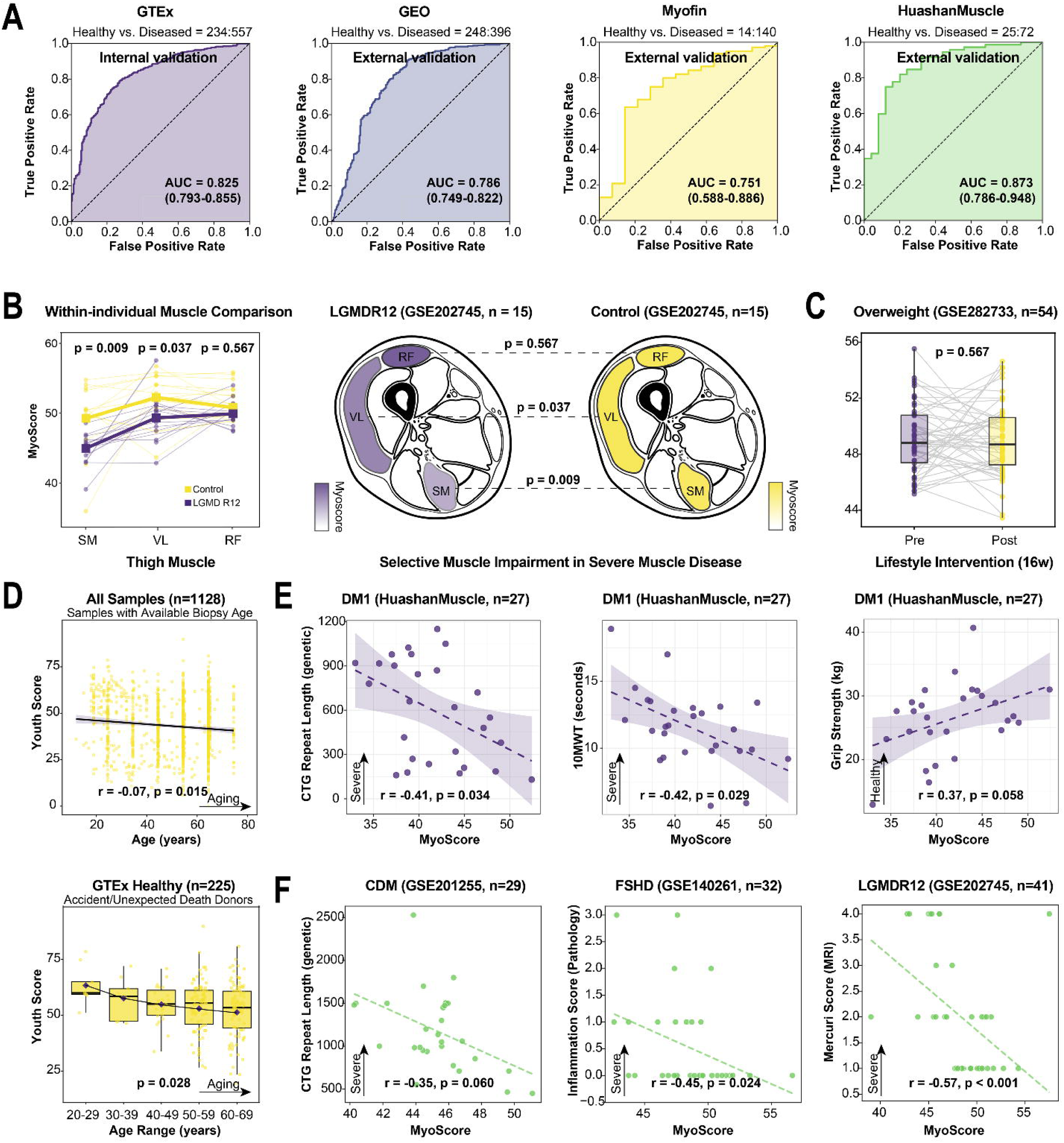
Disease discrimination and clinical validation. **a**, ROC curves for four cohorts: GTEx (AUC = 0.825), GEO (0.786), Myofin (0.751) and HuashanMuscle (0.873). Shaded areas indicate 95% CIs. **b**, Selective muscle impairment in LGMD R12: semimembranosus (P = 0.009), vastus lateralis (P = 0.037) and rectus femoris (P = 0.567). **c**, MyoScore stability after 16-week lifestyle intervention (n = 54; P = 0.567). **d**, Youth score versus chronological age (n = 1,128; r = −0.07, P = 0.015). **e**, Clinical correlations in DM1: CTG repeats (r = −0.41), 10-metre walk (r = −0.42) and grip strength (r = 0.37). **f**, Cross-disease correlations: CDM CTG repeats, FSHD inflammation and LGMD R12 Mercuri score.

Within individual patients, MyoScore captured disease-specific patterns of selective muscle involvement (Fig. 5B). In patients with LGMD R12 (anoctaminopathy due to recessive *ANO5* mutations), semimembranosus was most severely affected (P = 0.009 versus controls), vastus lateralis showed intermediate involvement (P = 0.037) and rectus femoris was relatively preserved (P = 0.567), consistent with the known posterior-predominant pattern of muscle involvement in anoctaminopathy^35^. MyoScore showed no significant change after a 16-week lifestyle intervention in overweight individuals (n = 54, P = 0.567) (Fig. 5C). This stability is consistent with MyoScore’s design as a measure of genetically regulated expression: because the scoring genes are identified through cis-eQTL effects, their basal expression levels retain a stable genetically regulated component and are less sensitive to acute environmental stimuli. Moreover, transcriptomic remodelling of skeletal muscle typically requires sustained, high-intensity stimuli over longer periods than the moderate lifestyle intervention studied here^36^. The Youth dimension correlated negatively with chronological age (r = −0.07, P = 0.015, n = 1,128) and declined progressively across decades in healthy GTEx donors (Kruskal–Wallis P = 0.028) (Fig. 5D).

Clinical correlations were examined across multiple disease cohorts (Fig. 5E, F; all correlations are Spearman’s rank correlation unless otherwise noted). In 27 patients with DM1, MyoScore correlated inversely with CTG repeat length (r = −0.41, P = 0.034), 10-metre walk time (r = −0.42, P = 0.029) and positively with grip strength (r = 0.37, P = 0.058). In CDM patients (n = 29), MyoScore trended inversely with CTG repeats (r = −0.35, P = 0.060). In FSHD patients (n = 32), MyoScore correlated with histological inflammation score (r = −0.45, P = 0.024). In LGMD R12 patients (n = 41), MyoScore showed strong inverse correlation with Mercuri MRI score (r = −0.57, P < 0.001).

### Histopathological validation reveals dimension-specific patterns

To validate MyoScore against gold-standard pathological assessment, we developed an automated quantification pipeline for haematoxylin and eosin (H&E)-stained whole slide images, yielding four metrics: nuclear centralization index, fibre size variability, fat infiltration percentage and fibrosis percentage (Fig. 6A, B). Critically, the H&E sections and RNA-seq data were derived from the same muscle biopsy specimen for each individual, enabling direct within-sample comparison between transcriptomic scores and histological features.

**Fig. 6.**
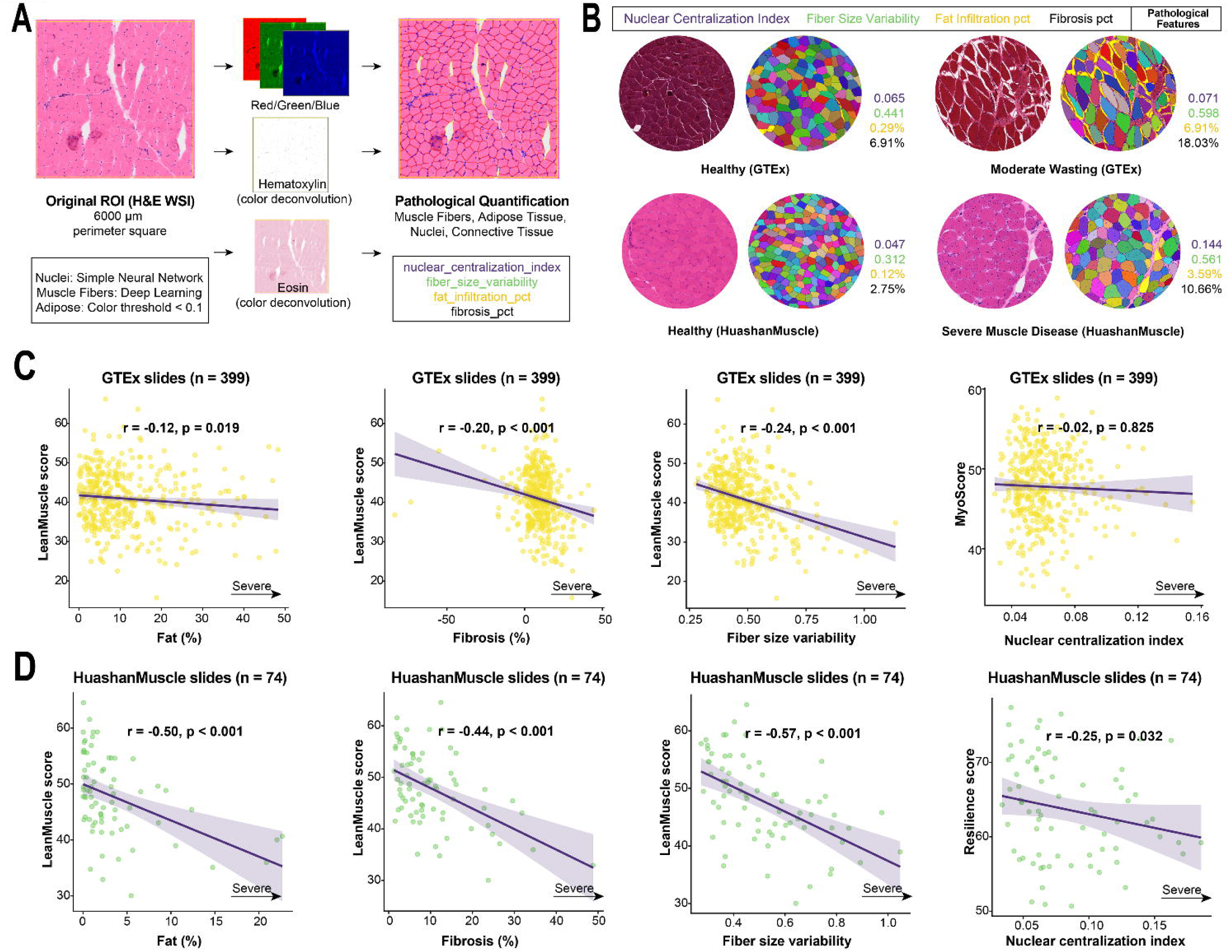
Histopathological validation. **a**, Automated H&E quantification pipeline. **b**, Representative images: healthy GTEx versus severe HuashanMuscle disease. **c**, Dimension–pathology correlations in GTEx (n = 399). **d**, Dimension–pathology correlations in HuashanMuscle disease cohort (n = 74). Spearman correlations; *P < 0.05, **P < 0.01, ***P < 0.001.

In the GTEx cohort (n = 399 slides), LeanMuscle showed the most consistent pathological correlations: negative associations with fat infiltration (r = −0.12, P = 0.019), fibrosis (r = −0.20, P < 0.001) and fibre size variability (r = −0.24, P < 0.001) (Fig. 6C). In the HuashanMuscle disease cohort (n = 74 slides), correlations were substantially stronger: LeanMuscle correlated negatively with fat infiltration (r = −0.50, P < 0.001), fibrosis (r = −0.44, P < 0.001) and fibre variability (r = −0.57, P < 0.001) (Fig. 6C). Resilience showed a significant negative correlation with nuclear centralization specifically in the disease cohort (r = −0.25, P = 0.032), suggesting that genetic resilience factors remain latent until challenged by pathological stress (Fig. 6D).

### MRI validation supports dimension-specific associations

Quantitative thigh MRI analysis from two independent cohorts confirmed dimension-specific associations at the within-individual level, where each patient’s transcriptomic MyoScore was compared with their own imaging data. In the HuashanMuscle cohort (n = 46), LeanMuscle correlated negatively with muscle fat fraction (r = −0.35, P = 0.018) and Mass correlated positively with muscle volume (r = 0.31, P = 0.037) (Fig. 7A, B). In the Myofin cohort (n = 13), the LeanMuscle–fat fraction correlation was even stronger (r = −0.62, P = 0.023), with Mass–volume showing a positive trend (r = 0.49, P = 0.089) (Fig. 7C, D). The replication of within-individual dimension-specific MRI correlations across two independent cohorts with different imaging protocols and ethnic backgrounds (Chinese and Finnish) confirms that TWAS-derived dimensions capture their intended biological constructs.

**Fig. 7.**
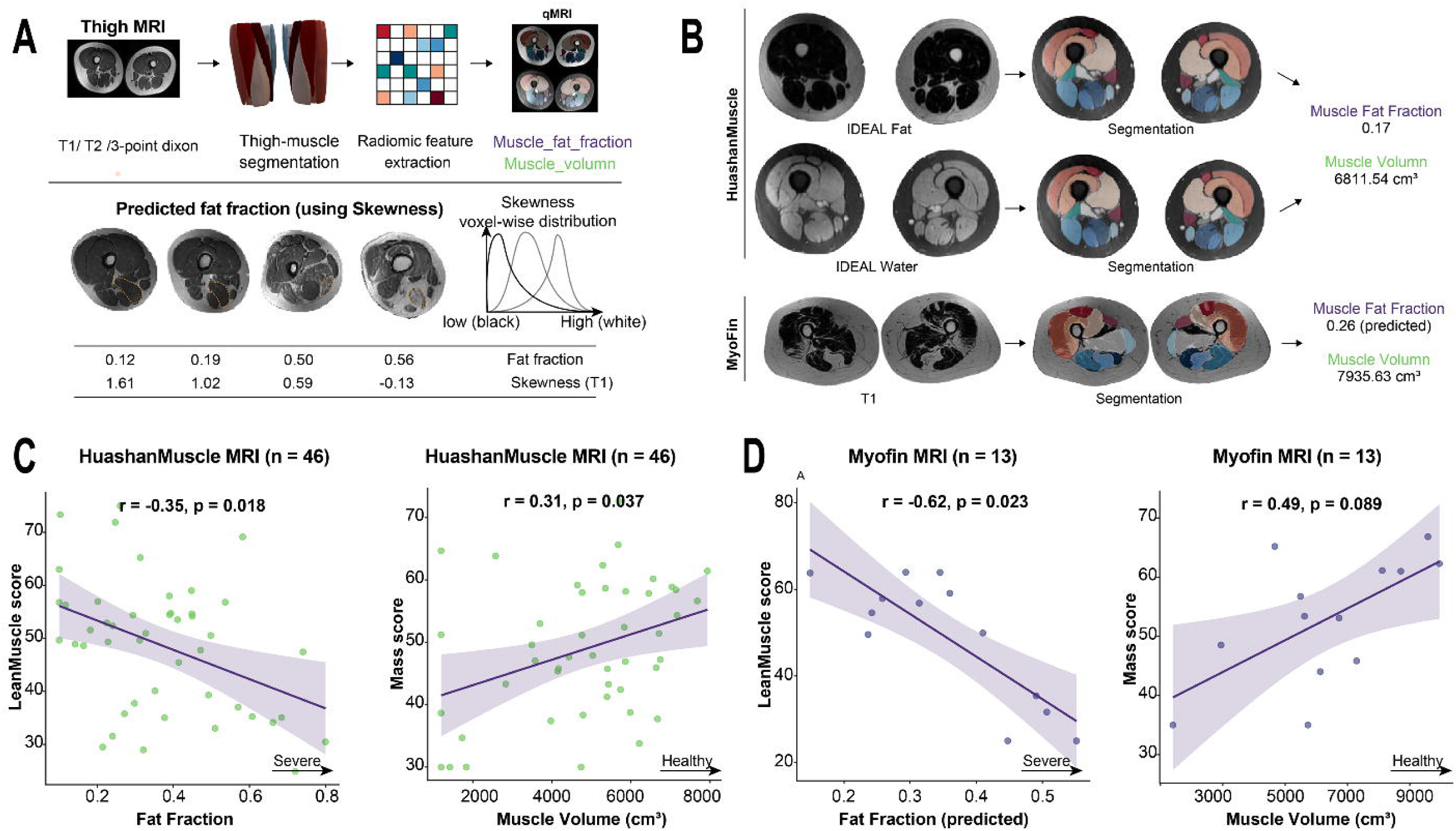
MRI validation of dimension-specific scores. **a,b**, HuashanMuscle cohort (n = 46): LeanMuscle versus fat fraction (r = −0.35, P = 0.018) and Mass versus volume (r = 0.31, P = 0.037). **c,d**, Myofin cohort (n = 13): LeanMuscle versus fat fraction (r = −0.62, P = 0.023) and Mass versus volume (r = 0.49, P = 0.087).

### Gene validation, causal inference and biomarker translation

The top 10 MyoScore genes, ranked by concordance × weight, were evaluated across 8 independent projects spanning 6 disease types (Fig. 8A). Three positive-direction genes (ACSS2^37^, TMEM52, GGT7^38^) and seven negative-direction genes (CEP250, UQCC1^39,40^, CPNE1^41,42^, RSRC2, RBL2^43–45^, SNRPC, GATAD1^46,47^) showed direction concordance rates of 50–88% across projects, with *TMEM52*, *RSRC2*, *CEP250* and *SNRPC* representing novel genes not previously associated with skeletal muscle function.

**Fig. 8.**
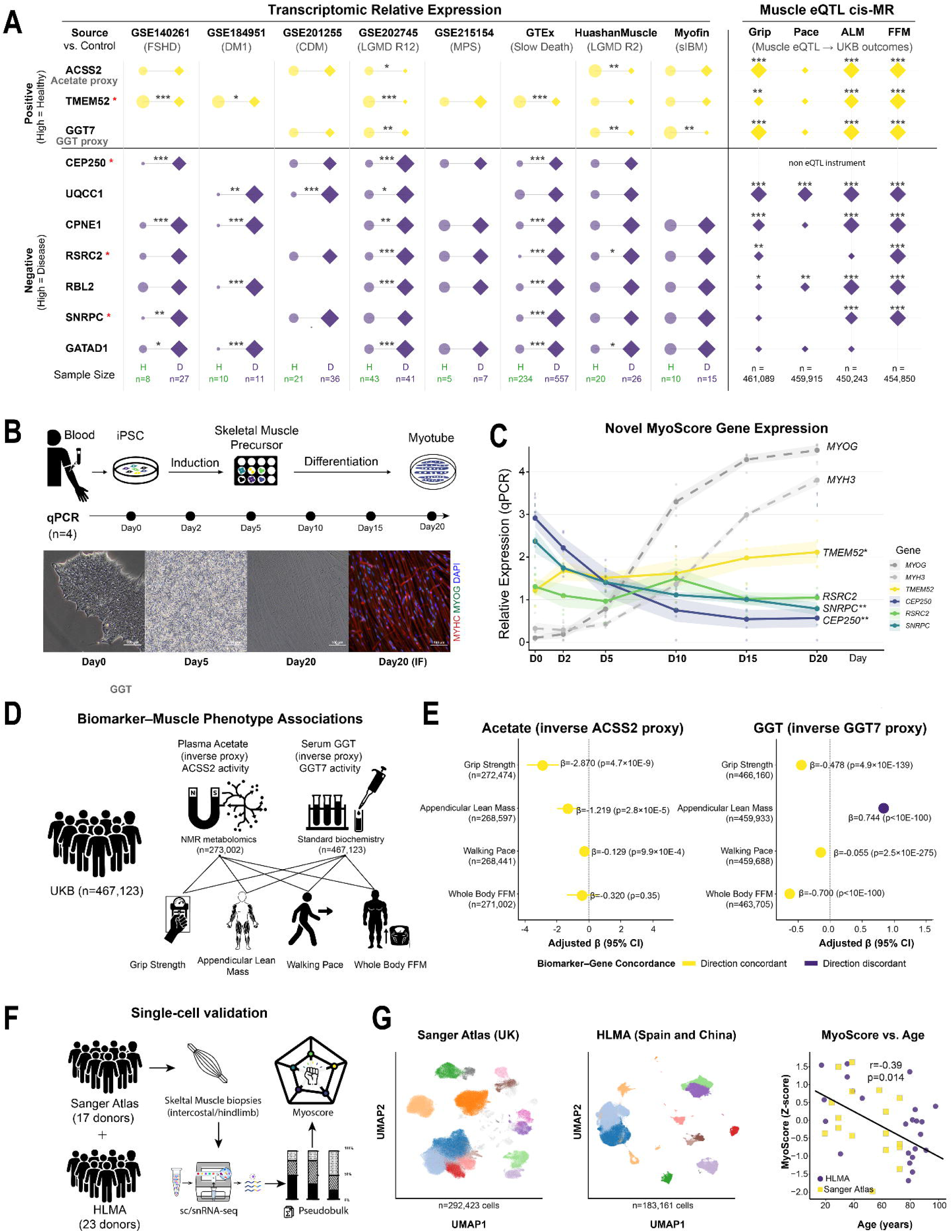
Novel gene validation, causal inference and single-cell validation. **a**, Left panel: cross-cohort transcriptomic concordance of top 10 MyoScore genes across 8 projects and 6 disease types. Each gene–cohort cell pairs a circle (H, healthy muscle) with a diamond (D, disease-related muscle); symbol area is proportional to mean expression, and per-group sample sizes are listed at the bottom. Right panel: two-sample Mendelian randomization using GTEx skeletal muscle cis-eQTL (n = 803) as instruments; overall concordance 78% (28/36 gene–outcome pairs). Yellow rows mark positive-direction genes (higher expression in healthy muscle); purple rows mark negative-direction genes (higher expression in diseased muscle). Genes flagged with a red asterisk (*) on the left are novel candidates without prior reports of skeletal-muscle relevance. **b**, iPSC-to-myotube differentiation time course for novel genes and myogenic markers (n = 4 donors, 6 time points). **c**, Gene expression fold changes (Day 20/Day 0) with significance from linear mixed models. **d**, UK Biobank biomarker–muscle phenotype associations schematic. **e**, Forest plots of adjusted β coefficients (95% CI) for plasma acetate and serum GGT associations with muscle phenotypes. **f**, Single-cell validation across two independent atlases. **g**, Pseudo bulk MyoScore versus donor age (n = 40 donors); combined Spearman ρ = −0.39, P = 0.014.

To test functional relevance, we tracked expression of four novel genes during iPSC-to-myotube differentiation in four healthy donors across six time points (Fig. 8B). Three of four genes showed statistically significant expression changes in directions consistent with MyoScore predictions (Fig. 8C): *TMEM52* showed progressive upregulation (fold change (FC) = 1.73, P = 3.6 × 10^−4^), while *CEP250* (FC = 0.20, P = 2.3 × 10^−7^) and *SNRPC* (FC = 0.33, P = 7.4 × 10^−7^) showed significant downregulation. *RSRC2* trended in the predicted direction (FC = 0.80) without reaching significance (P = 0.60). All four genes showed expression changes in the predicted direction (4/4 concordance).

To assess clinical relevance, we used circulating protein and biochemistry measurements in the UK Biobank as blood-based proxies for MyoScore gene products and tested their associations with health outcomes in up to 467,123 participants (Fig. 8D, E; per-outcome sample sizes varied). Plasma acetate was measured by NMR metabolomics in a subset of 273,002 participants, whereas serum GGT and the remaining biomarkers were assayed in the full clinical biochemistry panel (n = 467,123). Plasma acetate, the direct substrate of *ACSS2*, showed significant negative associations with grip strength (β = −2.870, P = 4.7 × 10^−9^, n = 272,474), appendicular lean mass (β = −1.219, P = 2.8 × 10^−5^, n = 268,597) and walking pace (β = −0.129, P = 9.9 × 10^−4^, n = 268,441), achieving 100% direction concordance across all muscle phenotypes. This is consistent with ACSS2’s role in acetate-to-acetyl-CoA conversion for muscle energy metabolism, where elevated plasma acetate reflects impaired clearance activity^37^. Serum gamma-glutamyl transferase (GGT; *GGT7* proxy, n = 467,123) similarly showed significant associations with grip strength (β = −0.478, P = 4.9 × 10^−139^) and walking pace (β = −0.055, P = 2.5 × 10^−275^), with 75% direction concordance^38^.

To support causal inference, we performed two-sample Mendelian randomization using skeletal muscle cis-eQTL from GTEx v8 (n = 803) as instruments and UK Biobank GWAS as outcomes (Fig. 8A, right panel). Across 9 genes with genome-wide significant cis-eQTL instruments (*CEP250* lacked a valid instrument), 28 of 36 gene–outcome pairs (78%) showed MR effect directions concordant with MyoScore predictions, with 27 reaching nominal significance (P < 0.05). *UQCC1* showed the strongest signal (appendicular lean mass P = 3.9 × 10^−243^), followed by CPNE1 (grip strength P = 5.6 × 10^−59^), *GGT7* (P = 4.5 × 10^−38^) and *ACSS2* (P = 1.6 × 10^−44^), each with 4/4 concordant outcomes. Notably, tissue specificity proved critical: when blood eQTL (eQTLGen) were used instead, *ACSS2* and *GGT7* showed fully discordant directions (0/4 concordant), resolved to 4/4 concordance with muscle eQTL, underscoring the importance of tissue-matched instruments for expression-trait MR.

### Single-cell validation across two independent atlases

To assess whether MyoScore, designed from bulk RNA-seq, retains biological validity at single-cell resolution, we computed MyoScore in two independent single-cell/single-nucleus RNA-seq atlases: the Human Limb Muscle Ageing atlas (HLMA; 292,423 cells from 23 donors aged 15–99 years)^48^ and the Sanger Skeletal Muscle atlas (183,161 cells from 17 donors aged 15–75 years)^49^ (Fig. 8F). All 417 MyoScore scoring genes were detected in both datasets.

Pseudobulk MyoScore (donor-level mean across all cell types) declined with chronological age in both atlases independently (HLMA: ρ = −0.34, P = 0.116; Sanger: ρ = −0.38, P = 0.135), and the combined cross-atlas correlation was significant (ρ = −0.39, P = 0.014, n = 40 donors; Fig. 8G). Among the five dimensions, Youth showed the strongest and most consistent ageing signal, declining in all 13 cell types examined in HLMA (Young > Old in every cell type). At the cell-type level, type II (fast-twitch) myofibre nuclei showed the largest MyoScore decline with age (Cohen’s d = 0.19, P = 1.5 × 10^−154^), followed by type I myofibre nuclei (d = 0.10) and myeloid cells (d = 0.13), consistent with the known preferential loss of type II fibres in ageing muscle^50^.

## Discussion

MyoScore represents a genetically anchored framework for objective quantification of muscle health across the complete spectrum from optimal function to severe pathology. By integrating TWAS results from 27 muscle-related phenotypes derived from over one million participants, we identified 1,116 TWAS-significant genes, of which 417 are expressed in skeletal muscle and organized into five biologically interpretable dimensions. Validation across 1,722 muscle transcriptomes from four independent cohorts demonstrates robust disease discrimination (AUC 0.751–0.873 across four cohorts), concordance with histopathological and imaging measures, and functional validation of genes previously uncharacterized in skeletal muscle biology.

The continuous muscle health spectrum revealed by transcriptomic embedding challenges the traditional binary classification of muscle health and disease. The significant alignment between MyoScore and the first diffusion component (r = 0.417) indicates that MyoScore captures the dominant axis of variation defined by its own 417-gene set under unsupervised embedding. The four-stage classification, spanning chronic illness, cancer cachexia and hereditary myopathies, provides a clinically intuitive framework bridging the gap between categorical diagnosis and the continuous reality of muscle health variation. This complements recent molecular subtyping of cancer cachexia muscle^51,52^, which identified disease-specific subtypes within the wasting continuum, while MyoScore offers a pan-disease, genetically anchored quantification across the full health spectrum.

The five dimensions capture fundamentally different aspects of muscle biology, reflected in their distinct pathological, imaging and ageing correlations. LeanMuscle showed the strongest histopathological correlations (r = −0.50 to −0.57 in the disease cohort) and MRI fat-fraction associations (r = −0.35 to −0.62), directly reflecting tissue composition. Mass tracked muscle volume on quantitative MRI across two independent cohorts (r = 0.31 to 0.49), consistent with its derivation from lean-mass and fat-free-mass GWAS phenotypes. Strength, anchored in grip-strength and walking-pace GWAS, showed the largest direction-concordant effects in UK Biobank plasma-acetate and cis-MR analyses, linking the dimension to acetyl-CoA–based energy metabolism. Youth, derived from telomere-length GWAS, showed the clearest age-related decline across bulk and single-cell data, with type II myofibre nuclei preferentially affected in the two muscle ageing atlases. Finally, the Resilience dimension showed context-dependent behaviour, with minimal correlation with pathology in healthy tissue but significant association with nuclear centralization in disease (r = −0.25), suggesting that genetic resilience factors remain latent until challenged by pathological stress, analogous to the concept of cognitive reserve in neurodegeneration^53,54^.

Pathway analyses further contextualize these dimensions in coherent metabolic and cellular programmes. The Strength dimension’s enrichment for acetyl-CoA metabolism positions MyoScore within the broader framework of skeletal muscle as a central metabolic organ^55^, where substrate utilization efficiency is a key determinant of health. The Youth dimension’s enrichment for mRNA methyltransferase activity connects biological ageing to epitranscriptomic regulation of S-adenosylmethionine homeostasis^29^, a pathway linked to age-related NAD+ decline and consistent with recent findings that trigonelline, an NAD+ precursor reduced in sarcopenia, improves muscle function in aged mice^56^. The Resilience dimension’s iron-sulfur cluster enrichment implicates ferroptosis susceptibility^32^ and mitochondrial quality control^57^ as determinants of disease resistance, complementing evidence that chaperone-mediated autophagy decline in skeletal muscle causes progressive myopathy^58^.

The functional validation of genes previously uncharacterized in skeletal muscle advances MyoScore beyond associative evidence at multiple levels. iPSC-to-myotube differentiation demonstrated that computationally identified genes show expression changes during myogenesis consistent with predictions in a disease-independent system. *CEP250*’s dramatic downregulation (FC = 0.20) during differentiation is biologically plausible, as this centrosomal protein essential for mitotic spindle formation declines during permanent cell cycle exit in myotube formation^59^. *TMEM52*’s progressive upregulation paralleling myogenic markers identifies a novel gene in muscle biology. The UK Biobank analysis extends these findings to population scale, establishing that *ACSS2*’s blood proxy (plasma acetate) shows 100% direction concordance across all muscle outcomes through a direct mechanistic link in muscle energy metabolism. Critically, two-sample Mendelian randomization using tissue-matched skeletal muscle cis-eQTL elevated these associations from observational to supporting causal evidence: 78% of gene–outcome pairs showed concordant MR directions, and the tissue specificity of instruments proved essential. *ACSS2* and *GGT7* showed fully discordant directions with blood eQTL but 4/4 concordance with muscle eQTL. This finding reinforces recent arguments for tissue-appropriate instruments in expression-trait MR^17^ and demonstrates that the direction assignments in MyoScore reflect genuine causal biology rather than confounded associations.

These findings have immediate translational applications. MyoScore offers an objective, quantitative measure for patient stratification in clinical trials, potentially reducing sample size requirements through more homogeneous cohort selection. Dimension-specific profiles could guide targeted therapeutic approaches: predominant Youth decline in ageing-related sarcopenia versus LeanMuscle deterioration in dystrophies. The identification of blood biomarker proxies (plasma acetate, serum GGT) opens possibilities for non-invasive monitoring, and the ability to calculate MyoScore from standard RNA-seq data enables retrospective analysis of existing cohorts.

Several limitations should be considered. The GWAS data underlying MyoScore derive predominantly from European-ancestry populations, and replication in more diverse cohorts will be needed as non-European muscle GWAS become available. Our validation is cross-sectional; longitudinal studies are needed to establish MyoScore’s utility for tracking disease progression and treatment response. The current implementation captures 417 of 1,116 identified genes, reflecting both appropriate exclusion of genes expressed primarily outside skeletal muscle and a residual subset below bulk RNA-seq detection that may be recoverable with single-cell or deeper-coverage sequencing. The UK Biobank analysis relies on circulating proxies whose informativeness depends on blood–tissue correspondence; the discordance between blood- and muscle-eQTL MR directions for ACSS2 and GGT7 underscores this limitation and motivated our use of tissue-matched instruments. Finally, the iPSC validation used four donors and cannot fully recapitulate adult muscle biology, and the cis-MR analysis relied on single-instrument Wald-ratio estimates without colocalization; multi-instrument MR with colocalization will strengthen causal attribution as additional eQTL signals and higher-powered outcome GWAS become available.

Future directions include deeper cell-type-resolved analysis to delineate cell-type-specific contributions to each dimension and identify the cellular origins of age-related score decline, multi-instrument cis-MR with colocalization analysis to further strengthen causal inference for the full gene set, and development of a simplified clinical assay targeting the most informative genes, particularly those with validated blood biomarkers, to enable routine clinical implementation without muscle biopsy.

In summary, MyoScore offers a genetically anchored, transcriptome-based framework for quantifying human skeletal muscle health along five biologically meaningful dimensions. By integrating TWAS-prioritized gene sets with muscle-specific expression data and orthogonal validation in independent cohorts, MyoScore moves beyond descriptive transcriptomics toward a mechanistically grounded, translationally oriented composite score. Although further prospective validation and assay simplification are required before clinical deployment, MyoScore provides a reproducible, interpretable foundation on which future multi-omic, longitudinal, and interventional studies of human muscle health can be built.

## Methods

### GWAS data collection and processing

We obtained summary statistics for 27 muscle-related phenotypes from publicly available GWAS studies (Supplementary Table 1). Primary data sources included the OpenGWAS database (IEU), UK Biobank summary statistics (Neale Lab and UK Biobank official releases) and FinnGen release 9. Combined GWAS sample sizes exceeded 1,000,000 participants of mixed ancestry. All summary statistics were harmonized to GRCh37/hg19 coordinates and filtered for minor allele frequency > 1% and imputation quality score > 0.8.

### TWAS analysis

Transcriptome-wide association analysis was performed using FUSION v1.1.0^16^. Pre-computed expression weights for skeletal muscle from GTEx v8 comprised 2,975 genes with significant cis-heritability, derived from 803 muscle samples using multiple prediction models (BLUP, BSLMM, LASSO, elastic net, top1 SNP), with the best-performing model selected per gene on the basis of cross-validation R^2^. The 1000 Genomes Phase 3 European ancestry reference panel (n = 503) provided linkage disequilibrium estimates. Results were concatenated across chromosomes with Bonferroni correction (0.05/2,975 = 1.68 × 10^−5^)

### Gene assignment and dimension construction

Genes achieving Bonferroni significance in at least one phenotype were assigned to five dimensions on the basis of biological categorization: Strength (grip strength, walking pace, muscle weakness, grip cross-sectional area; 5 phenotypes), Mass (15 lean/fat-free mass phenotypes), LeanMuscle (anterior/posterior thigh fat infiltration; 2 phenotypes, direction reversed), Youth (telomere length; 1 phenotype) and Resilience (muscular dystrophy, myopathies, creatine kinase; 5 phenotypes, direction reversed). Gene weights were calculated as −log^10^(TWAS P value). Effect direction was determined from TWAS Z-scores with reversals applied to ensure consistent interpretation (higher score = better health).

### Ethics

This study was approved by the relevant institutional review boards. The Helsinki Myofin cohort was approved by the Ethics Committee of Helsinki University Hospital (HUS; approval number 195/13/03/00/11), and written informed consent was obtained from each participant. The HuashanMuscle cohort was collected at Huashan Hospital, Fudan University, under institutional review board approval (approval numbers 2022-913 and 2019-409), and written informed consent was obtained from all participants. iPSC lines were generated from peripheral blood mononuclear cells of four healthy donors who provided written informed consent under the same Huashan Hospital ethical approvals. The GTEx dataset was accessed through the Genotype-Tissue Expression Project (dbGaP accession phs000424); all GTEx tissue samples were collected from deceased donors under informed consent from next of kin. UK Biobank data were accessed under approved application number 19542. GEO datasets were obtained from publicly available repositories; ethics approvals for these cohorts were obtained by the original investigators as described in the respective publications. All procedures conformed to the principles of the Declaration of Helsinki.

### RNA-seq data processing

This study extends our previously established myopathy spectrum framework^9^, which integrated transcriptional and clinical features of human skeletal muscles across varying health conditions. Building on that foundation, we aggregated 1,722 human skeletal muscle RNA-seq samples (Supplementary Table 2) from four sources: GTEx v8 (n = 803)^17^, GEO datasets (n = 668, 15 studies)^36,52,60–70^, Helsinki Myofin (n = 154)^9,71^ and Shanghai HuashanMuscle (n = 97). For inclusion, datasets were required to meet the following criteria: (1) human skeletal muscle tissue (no cell lines or organoids); (2) bulk RNA sequencing performed using high-throughput platforms (excluding microarrays and single-cell data); and (3) data available in raw count format (datasets shared only in transformed count format were excluded). Raw count data were normalized using trimmed mean of M-values (TMM) via edgeR^72^, and log_2_(CPM + 1) transformation was applied for score calculation.

### MyoScore calculation

MyoScore was calculated in five steps (Supplementary Note 1, Supplementary Table 2): (1) selection of dimension-specific genes present in expression data; (2) gene-wise Z-score normalization across all samples; (3) weighted averaging within each dimension: dimension score = Σ(z-score × direction × weight)/Σ(weight); (4) min–max scaling of each dimension score to 0–100 using the current 1,722-sample reference distribution; and (5) composite scoring: MyoScore = 0.252 × Strength + 0.177 × Mass + 0.243 × LeanMuscle + 0.242 × Youth + 0.087 × Resilience. The composite weights α_d were derived on the GTEx v8 skeletal muscle reference cohort (n = 803) as the normalized absolute Spearman rank correlation between each dimension sub-score and the first principal component of the five sub-scores within GTEx; this keeps weight derivation independent of the external disease cohorts used for validation (Supplementary Table 3). UMAP dimensionality reduction^73^ was performed on the batch-corrected, log^2^(CPM + 1)-transformed expression matrix using MyoScore gene sets (n_neighbors = 15, min_dist = 0.1). Diffusion components were computed using the diffusion map algorithm^74^ on the same expression matrix.

### Pathway enrichment analysis

Gene ontology and KEGG pathway enrichment analyses were performed using Enrichr^75^ via the GSEApy Python package^76^(Supplementary Table 4). For each dimension, protein-coding genes were extracted (excluding non-coding RNAs) and tested against GO Biological Process 2023, GO Molecular Function 2023 and KEGG 2021 Human gene set libraries. Analyses were performed on three gene sets per dimension: all genes, healthy-direction genes (direction = +1) and unhealthy-direction genes (direction = −1). Significance was defined as Benjamini–Hochberg adjusted P < 0.05.

### Histopathology quantification

H&E-stained muscle sections were digitized as whole slide images (Supplementary Table 5). Quantitative analysis selected 6,000-μm perimeter-square regions of interest and applied colour deconvolution, neural network-based nuclei detection, deep learning-based muscle fibre segmentation and colour thresholding for adipose tissue identification. Four pathological features were quantified: fat infiltration percentage, fibrosis (collagen area fraction), fibre size variability (coefficient of variation in cross-sectional area) and nuclear centralization index. Analysis was performed on 399 GTEx slides and 74 HuashanMuscle slides (https://github.com/Hirriririir/MyoPath).

### Thigh MRI quantification

Thigh MRI data were acquired from HuashanMuscle (n = 46) and Myofin (n = 13) cohorts (Supplementary Table 6). For HuashanMuscle, IDEAL fat and water images were used to derive muscle fat fraction and volume. For the Myofin cohort lacking Dixon^77^ sequences, fat fraction was predicted from T1-weighted images using voxel-wise signal distribution skewness (https://github.com/Hirriririir/Multimodal-Multiethnic-Thigh-Muscle-MRI-analysis).

### iPSC differentiation and qPCR

Four novel MyoScore genes (*TMEM52*, *CEP250*, *RSRC2*, *SNRPC*) were selected on the basis of novelty and high model weight, with *MYOG* and *MYH3* serving as myogenic positive controls. iPSC lines from four healthy donors were established from peripheral blood mononuclear cells by Shanghai Amplicell Biosciences (Shanghai, China) using a non-integrating Sendai virus (SeV) reprogramming approach, in which temperature-sensitive vector mutations enable transgene-free iPSC generation and efficient viral clearance at non-permissive temperatures^78^. Quality control included morphological assessment, mycoplasma testing, pluripotency marker immunofluorescence, and SeV clearance confirmed by RT-PCR and anti-SeV immunostaining. iPSC lines were subsequently differentiated into skeletal myotubes and RNA was collected at six time points (Day 0, 2, 5, 10, 15, 20)^79^; qPCR was normalized to GAPDH and ACTB (Supplementary Table 7).

### UK Biobank biomarker analysis

Plasma acetate (*ACSS2* proxy; NMR metabolomics, n = 273,002) and serum GGT (*GGT7* proxy; standard biochemistry, n = 467,123) were tested for association with grip strength, appendicular lean mass, walking pace and whole-body fat-free mass using ordinary least squares regression with heteroscedasticity-consistent standard errors (HC3), adjusting for age, sex, body mass index, ethnicity and assessment centre. GGT was log-transformed (Supplementary Table 8).

### Two-sample Mendelian randomization

Two-sample MR was performed using the TwoSampleMR R package (v0.7.0). For cis-MR of MyoScore genes, genome-wide significant cis-eQTL (P < 5 × 10^−8^) for each gene were obtained from GTEx v8 skeletal muscle (n = 803) via the eQTLGen and GTEx portals. The top cis-eQTL per gene was used as a single instrument, and causal effects were estimated using Wald ratio. Outcomes were UK Biobank GWAS for grip strength (ukb-b-10215, n = 461,089), walking pace (ukb-b-4711, n = 459,915), appendicular lean mass (ebi-a-GCST90000025, n = 450,243) and whole-body fat-free mass (ukb-b-13354, n = 454,850). Direction concordance was defined as the MR effect direction matching the MyoScore gene direction assignment (positive-direction genes: higher expression → better muscle outcome; negative-direction genes: higher expression → worse outcome). For comparison, analyses were repeated with blood eQTL from eQTLGen (n = 31,684). *CEP250* was excluded from cis-MR owing to the absence of a genome-wide significant cis-eQTL instrument (Supplementary Table 9).

### Single-cell MyoScore analysis

MyoScore was computed in two independent single-cell/single-nucleus RNA-seq atlases. The HLMA atlas^48^ comprised 292,423 cells/nuclei from 23 donors (aged 15–99 years; sc/snRNA-seq), with pre-computed UMAP embeddings and cell type annotations for 15 cell types. The Sanger Skeletal Muscle atlas^49^ comprised 183,161 cells/nuclei from 17 donors (aged 15–75 years; sc/snRNA-seq), with annotations at three hierarchy levels (up to 83 subtypes). For each cell, per-gene expression was Z-score-normalized across all cells, dimension scores were computed as weighted averages (using the same gene weights as bulk MyoScore), and a composite MyoScore was calculated using standard dimension weights. Pseudobulk scores were obtained by averaging MyoScore across all cells per donor. To combine donors from both atlases on a common scale, within-atlas Z-score normalization was applied before cross-atlas correlation analysis. Age–MyoScore associations were assessed using Spearman’s rank correlation. Young–old comparisons at the cell-type level used Mann–Whitney U tests with Bonferroni correction, and effect sizes were quantified as Cohen’s d (Supplementary Table 10).

### Statistical analysis

Disease discrimination was assessed using ROC analysis (pROC R package^80^). AUC and 95% CIs were calculated using DeLong’s method. Correlations used Spearman’s rank correlation. Partial correlations controlling for confounders used ppcor^81^. Group comparisons used Wilcoxon rank-sum tests (unpaired) or Wilcoxon signed-rank tests (paired). iPSC gene expression was analysed using linear mixed models (expression ∼ day + (1|donor)). UK Biobank analyses used Benjamini–Hochberg false discovery rate correction. Statistical significance was set at P < 0.05 unless otherwise noted. All tests were two-sided.

## Data Availability

The integrated and processed RNA-seq datasets, MyoScore results, gene weights and calculation scripts used in this study are available at https://github.com/Hirriririir/MyoScore. The following public data sources were used in this study:

### Transcriptomic data

GTEx v8 skeletal muscle RNA-seq (n = 803) is available from the GTEx Portal (gtexportal.org). GEO datasets are publicly available at ncbi.nlm.nih.gov/geo (accession numbers listed in Supplementary Table 2).

### GWAS summary statistics

GWAS data for 27 muscle-related phenotypes were obtained from the OpenGWAS database (gwas.mrcieu.ac.uk; IEU identifiers listed in Supplementary Table 1), the UK Biobank Neale Lab round 2 (nealelab.is/uk-biobank) and FinnGen release 9 (finngen.gitbook.io/documentation).

### eQTL and TWAS resources

GTEx v8 skeletal muscle cis-eQTL summary statistics and pre-computed FUSION TWAS weights are available from the GTEx Portal and the FUSION TWAS website (gusevlab.org/projects/fusion). The 1000 Genomes Phase 3 European LD reference panel is available from the International Genome Sample Resource (internationalgenome.org). Blood eQTL data from eQTLGen (n = 31,684) are available at eqtlgen.org.

### Single-cell/single-nucleus RNA-seq atlases

The HLMA (Human Limb Muscle Ageing) atlas (292,423 cells/nuclei, 23 donors) is available from the CNGB database (db.cngb.org/cdcp/hlma). The Sanger Skeletal Muscle atlas is available from the CellxGene portal (cellxgene.cziscience.com).

### UK Biobank

Individual-level data for biomarker analyses (plasma acetate, serum GGT, grip strength, appendicular lean mass, walking pace, whole-body fat-free mass) were accessed under application 19542.

### Histopathological data

H&E whole slide images from the HuashanMuscle cohort, together with processed GTEx slides, are publicly accessible via the MyoToolkit portal (myotoolkit.huashanmuscle.com); the GTEx slides are additionally available from the GTEx Portal (gtexportal.org).

Other data not publicly deposited are available from the corresponding authors upon reasonable request.

## Code Availability

The MyoScore R package, including TWAS integration, dimension scoring, and all statistical analyses described in this study, is available at https://github.com/Hirriririir/MyoScore. An interactive web-based MyoScore calculator is available at https://myotoolkit.huashanmuscle.com/myoscore/calculator.html. Automated quantification of H&E-stained whole slide images, encompassing colour deconvolution, neural network-based nuclei detection, deep learning muscle fibre segmentation and adipose/fibrosis thresholding, was performed using MyoPath, available at https://github.com/Hirriririir/MyoPath. Thigh MRI fat fraction and muscle volume quantification, including the T1-based fat fraction prediction pipeline for cohorts lacking Dixon sequences, was performed using code available at https://github.com/Hirriririir/Multimodal-Multiethnic-Thigh-Muscle-MRI-analysis.

## Acknowledgements

We thank the participants and their families who donated their muscle tissues for research purposes. We would also like to extend special thanks to the authors of these publicly available muscle datasets, which will facilitate further research in the future.

## Author Contributions

H.Z. conceived and designed the study, developed the MyoScore framework, performed the analyses and wrote the manuscript. S.Ma. contributed to data curation, histological experiments, single-cell analyses and manuscript drafting. V.L. curated the Helsinki Myofin cohort, benchmarked the MyoScore R package and reviewed the manuscript. M.G. and W.Z. performed histological experiments. M.J. contributed samples and clinical data and provided methodological advice on statistics and causal inference. A.O. provided bioinformatics input and manuscript review. P.H.J. provided editorial review and scientific advice. P.H. provided scientific advice. J.X. and W.Zh. collected and phenotyped Huashan cohort samples. Sh.Z. provided computational resources. B.U. and C.Z. provided cohort resources and scientific advice. M.S. and S.L. conceived and supervised the study and provided cohort resources. All authors read and approved the final manuscript.

## Supplementary Information

**Supplementary Note 1.** Mathematical Framework of MyoScore.

**Supplementary Table 1**. GWAS data sources for MyoScore construction. Summary of 27 genome-wide association studies spanning five MyoScore dimensions.

**Supplementary Table 2**. Metadata for 1,722 human skeletal muscle RNA-seq samples.

**Supplementary Table 3**. Sample characteristics and MyoScore distributions by disease phenotype.

**Supplementary Table 4**. Pathway enrichment results for MyoScore gene sets across five dimensions (GO Biological Process, GO Molecular Function, and KEGG).

**Supplementary Table 5**. Histopathology quantification results of 399 GTEx slides and 74 HuashanMuscle slides.

**Supplementary Table 6**. MRI results of 46 HuashanMuscle individuals and 13 Myofin individuals.

**Supplementary Table 7**. iPSC qPCR data of 4 HuashanMuscle donors.

**Supplementary Table 8**. UK Biobank biomarker analysis for *ACSS2* and *GGT7* proxies.

**Supplementary Table 9**. Cis-Mendelian randomization results for top MyoScore genes using skeletal muscle eQTL.

**Supplementary Table 10**. Single-cell MyoScore analysis results for the Sanger Skeletal and the HLMA datasets.

