## Supplementary File 1 for "MyoScore: a Genetically Anchored Transcriptomic Scoring System for Quantifying Human Skeletal Muscle Health"

**Supplementary Note 1.** Mathematical Framework of MyoScore.

**Supplementary Note 1**

**Mathematical Framework of MyoScore**

**1. Gene selection and weight assignment**

**1.1 TWAS gene identification**

For each of the 27 muscle-related GWAS phenotypes, transcriptome-wide association analysis was performed using FUSION. A gene *g* was considered significant for phenotype *p* if its TWAS P value passed Bonferroni correction:

*Pg,p < 0.05 / Ngenes = 0.05 / 2,975 = 1.68 × 10⁻⁵*

where *N*genes = 2,975 is the number of genes with significant cis-heritability in GTEx v8 skeletal muscle.

**1.2 Dimension assignment**

Each significant gene was assigned to one of five dimensions (*d* ∈ {Strength, Mass, LeanMuscle, Youth, Resilience}) based on the biological categorization of its source GWAS phenotype(s). Genes significant in multiple phenotypes within the same dimension were retained with aggregated statistics.

**1.3 Gene weight**

The weight of gene *g* in dimension *d* was defined as:

*wg,d = −log₁₀(Pg,d)*

where *P*g,d is the minimum TWAS P value for gene *g* across all phenotypes assigned to dimension *d*. Higher statistical significance yields greater weight in the scoring.

**1.4 Direction assignment**

The effect direction of gene *g* was determined from its TWAS Z-score:

*δg = +1 if Zg > 0 for positive phenotypes (e.g., grip strength)*

*δg = −1 if Zg < 0 for positive phenotypes*

For negative phenotypes (where higher values indicate worse health, such as thigh fat infiltration, creatine kinase and myopathy diagnoses), the direction was inverted:

*δgᵢⁿᵃˡ = −δg*

This ensures a consistent interpretation across all dimensions: *δ*gᵢⁿᵃˡ = +1 means higher expression of gene *g* indicates better muscle health.

**2. MyoScore calculation**

**2.1 Expression preprocessing**

Given a raw count matrix **X** ∈ ℝG×N (genes × samples), the expression was normalized as follows.

***Step 1: Counts per million (CPM)***

*CPMg,s = ( Xg,s / Σg'=1…G Xg',s ) × 10⁶*

***Step 2: Log transformation***

*Eg,s = log₂(CPMg,s + 1)*

**2.2 Gene-wise Z-score normalization**

For each scoring gene *g*, expression was standardized across all *N* samples:

*zg,s = (Eg,s − Ēg) / σg*

where *Ē*g = (1/N) Σs=1…N Eg,s and σg = √[ (1/(N−1)) Σs=1…N (Eg,s − Ēg)² ].

**2.3 Dimension sub-score**

For each dimension *d*, the raw score of sample *s* was computed as a direction- and weight-adjusted average:

*Sʳᴀʷg,s = (Σg∈Gd zg,s · δg · wg,d) / (Σg∈Gd wg,d)*

where *G*d is the set of genes assigned to dimension *d* that are detectable in the expression data. The raw score was then rescaled to [0, 100] via min-max normalization across all samples:

*Sd,s = [ Sʳᴀʷd,s − min(Sʳᴀʷd,s) ] / [ max(Sʳᴀʷd,s) − min(Sʳᴀʷd,s) ] × 100*

**2.4 Composite MyoScore**

The five dimension sub-scores were combined into a composite MyoScore using data-driven weights:

*MyoScores = Σd=1…5 αd · Sd,s*

where the dimension weights *α*d were:

| **Dimension** | **Symbol** | **Weight (αd)** |
| --- | --- | --- |
| *Strength* | α₁ | 0.252 |
| *Mass* | α₂ | 0.177 |
| *LeanMuscle* | α₃ | 0.243 |
| *Youth* | α₄ | 0.242 |
| *Resilience* | α₅ | 0.087 |

Explicitly:

*MyoScore = 0.252 × SStrength + 0.177 × SMass + 0.243 × SLeanMuscle + 0.242 × SYouth + 0.087 × SResilience*

**2.5 Data-driven weight derivation**

The five composite dimension weights *α_d_* (*d* ∈ {Strength, Mass, LeanMuscle, Youth, Resilience}) were derived on the 803 GTEx v8 skeletal muscle reference samples only. This keeps weight derivation within the same genetically characterised reference population that also provides the TWAS cis-eQTL weights, gene-direction assignments and gene-level weights used earlier in the framework, and independent of the external disease cohorts (GEO, Helsinki Myofin, Huashan) and single-cell ageing atlases that are subsequently used for validation.

For each GTEx sample we computed the five dimension sub-scores *S_d_* (as in §2.3) and summarised them with principal component analysis. The first principal component (PC1) explained 62.7% of variance, with all five dimension loadings of the same sign (all positive in the “higher = healthier” direction); PC1 was therefore taken as an unsupervised GTEx-internal muscle-health axis, defined without reference to any disease label in the external cohorts.

Each dimension weight was then set proportional to the absolute Spearman rank correlation between its sub-score and this GTEx-internal axis:

*α_d_ = |ρ(S_d_, PC1_GTEx_)| / Σ_d′=1…5_ |ρ(S_d′_, PC1_GTEx_)|*

yielding *α_Strength_* = 0.252, *α_Mass_* = 0.177, *α_LeanMuscle_* = 0.243, *α_Youth_* = 0.242 and *α_Resilience_* = 0.087.

Because PC1 is learned on GTEx samples only, neither the gene-direction assignment, the gene weight, nor the composite weight derivation is informed by the external disease cohorts or single-cell ageing atlases that serve as independent validations in the main text.

**2.6 Gene overlap and pleiotropy handling**

The 1,116 TWAS-significant associations correspond to 775 unique genes, of which 510 were significant in GWAS phenotypes mapping to a single MyoScore dimension, 206 to two dimensions, 43 to three, 15 to four, and one (*SULT1A2*) to all five dimensions. Restricting to genes detectable in bulk RNA-seq yields 591 gene-dimension entries from 417 unique genes.

Dimension sub-scores are computed independently over each dimension’s gene list (§2.3). A gene that is TWAS-significant on *N* dimensions therefore contributes to *N* dimension sub-scores, which after min-max normalisation (§2.3) enter the composite score with fixed weights *α_d_* (§2.5). This treatment reflects biological pleiotropy: a single gene can influence multiple muscle-health traits (e.g., grip strength and telomere length) through distinct physiological pathways, and is not an inadvertent double counting, as each (gene, dimension) association is independently supported by TWAS evidence on its assigned phenotype(s).

**3. Summary of key notation**

| **Symbol** | **Definition** |
| --- | --- |
| *g* | Gene index |
| *s* | Sample index |
| *d* | Dimension index (1–5) |
| *Pg,d* | TWAS P value for gene g in dimension d |
| *wg,d* | Gene weight: −log10(Pg,d) |
| *δg* | Direction (+1 = healthy, −1 = disease) |
| *zg,s* | Z-score-normalized expression |
| *Sd,s* | Dimension sub-score (0–100) |
| *αd* | Dimension weight (data-driven) |
| *MyoScores* | Composite score for sample s |
| *Gd* | Set of detectable genes in dimension d |

**4. Gene counts by dimension**

| **Dimension** | **Total TWAS genes** | **Detectable in bulk RNA-seq** | **Source phenotypes** |
| --- | --- | --- | --- |
| Strength | 67 | 31 | 5 |
| Mass | 382 | 219 | 15 |
| LeanMuscle | 272 | 147 | 2 |
| Youth | 81 | 37 | 1 |
| Resilience | 314 | 157 | 5 |
| ***Total*** | **1,116** | **591 entries (417 unique)** | **28** |

*Note: Some genes appear in multiple dimensions, yielding 591 gene-dimension entries from 417 unique genes.*
